# Protein-truncating variants in *BSN* are associated with severe adult-onset obesity, type 2 diabetes and fatty liver disease

**DOI:** 10.1101/2023.06.14.23291368

**Authors:** Yajie Zhao, Maria Chukanova, Katherine A Kentistou, Zammy Fairhurst-Hunter, Anna Maria Siegert, Raina Jia, Georgina Dowsett, Eugene J Gardner, Felix R Day, Lena R Kaisinger, Yi-Chun Loraine Tung, Brian Yee Hong Lam, Hsiao-Jou Cortina Chen, Quanli Wang, Jaime Berumen-Campos, Pablo Kuri-Morales, Roberto Tapia-Conyer, Jesus Alegre-Diaz, Jonathan Emberson, Jason M Torres, Rory Collins, Danish Saleheen, Katherine R Smith, Dirk S Paul, Florian Merkle, Nick J Wareham, Slavé Petrovski, Steve O’Rahilly, Ken K Ong, Giles S H Yeo, John R B Perry

## Abstract

Obesity is a major risk factor for many common diseases and has a significant heritable component. While clinical and large-scale population studies have identified several genes harbouring rare alleles with large effects on obesity risk, there are likely many unknown genes with highly penetrant effects remaining. To this end, we performed whole exome-sequence analyses for adult body mass index (BMI) in up to 587,027 individuals. We identified rare, loss of function variants in two genes – *BSN* and *APBA1* – with effects on BMI substantially larger than well-established obesity genes such as *MC4R*. One in ∼6500 individuals carry a heterozygous protein truncating variant (PTV) in *BSN*, which confers a 6.6, 3.7 and 3-fold higher risk of severe obesity (BMI >40kg/m^2^), non-alcoholic fatty liver disease and type 2 diabetes, respectively. In contrast to most other obesity-related genes, rare variants in *BSN* and *APBA1* had no apparent effect on childhood adiposity. Furthermore, *BSN* PTVs magnified the influence of common genetic variants associated with BMI, with a common polygenic score exhibiting an effect on BMI twice as large in *BSN* PTV carriers than non-carriers. Finally, we explored the plasma proteomic signatures of *BSN* PTV carriers as well as the functional consequences of *BSN* deletion in human iPSC-derived hypothalamic neurons. These approaches highlighted a network of differentially expressed genes that were collectively enriched for genomic regions associated with BMI, and suggest a role for degenerative neuronal synaptic function and neurotransmitter release in the etiology of obesity.

## Introduction

Over one billion people worldwide live with obesity, a global health challenge that is rapidly increasing in scale^1, 2^. Obesity is the second leading cause of preventable death, increasing the risks of diseases such as type 2 diabetes, cardiovascular disease and cancer^1, 3^. Understanding the full range of social, psychological, and biological determinants of energy intake and expenditure will be key to tackling this epidemic.

Early studies in mice highlighted the role of the leptin-melanocortin pathway in appetite and body weight regulation^4^, which led to candidate gene sequencing studies of rare individuals with severe early-onset obesity. Those studies identified rare loss of function mutations in key components of this pathway as causes of severe, early-onset obesity^5^, the most common of which impact the melanocortin 4 receptor (*MC4R*)^6, 7^. In parallel, using a ‘hypothesis-free’ approach, large-scale population-based genome-wide association studies (GWAS) have identified hundreds of common genetic variants associated with body mass index (BMI) in adults^8^. Those variants are mostly non-coding and are enriched near genes expressed in the brain^9^. Individually, the effect of each variant is small, and cumulatively the ∼1000 common variants identified to date explain only ∼6% of the population variance in BMI^8^. The recent emergence of whole exome sequence (WES) data at the population scale has enabled exome-wide association studies (ExWAS), leading to a convergence of common and rare variant discoveries. In a landmark study, Akbari *et al* used WES data in ∼640,000 individuals to identify rare protein-coding variants in 16 genes associated with BMI^10^. These included genes with established roles in weight regulation (*MC4R*, *GIPR* and *PCSK1*) in addition to novel targets, such as *GPR75*, in which loss-of-function mutations are protective against obesity in humans and mice^10^.

The current study is an ExWAS for BMI using WES data of 419,668 UK Biobank participants. Although this represents a subset of the exomes previously reported by Akbari *et al*^10^, we were motivated by recent work demonstrating that in the context of gene-burden analysis^11^, the various choices around how one could define a qualifying rare variant can highlight biologically relevant genes at exome-wide significance missed using alternative definitions^12^. Consistent with this, our approach identified novel rare variant associations with *BSN* and *APBA1*, which we replicated in independent WES data from 167,359 individuals of non-European genetic ancestries. The detected rare protein truncating variants in *BSN* and *APBA1* have larger effects than other previously reported ExWAS genes^10^, and our findings collectively suggest an emerging role for degenerative neuronal synaptic function and neurotransmitter release in the etiology of obesity.

## Results

To identify rare variants associated with adult BMI, we performed an ExWAS using genotype and phenotype data from 419,668 individuals of European ancestry from the UK Biobank study^13^. Individual gene-burden tests were performed by collapsing rare (MAF<0.1%) genetic variants across 18,658 protein-coding genes. We tested three categories of variants based on their predicted functional impact: high-confidence Protein Truncating Variants (PTVs), and two overlapping missense masks that used a REVEL^14^ score threshold of 0.5 or 0.7. This yielded a total of 37,691 gene tests with at least 30 informative rare allele carriers, corresponding to a multiple-test corrected statistical significance threshold of *P*<1.33×10^-6^ (0.05/37,691).

Genetic association testing was performed using BOLT-LMM^15^, which identified a total of nine genes meeting this threshold for significant association with adult BMI **(Table S1)**. Our gene-burden ExWAS appeared statistically well-calibrated, as indicated by low exome-wide test statistic inflation [*λ_GC_*=1.05-1.15] and by the absence of significant associations with any synonymous variant masks **(Figure S1-2)**. Five of our identified associations were previously reported – PTVs in *MC4R*, *UBR2*, *KIAA1109*, *SLTM* and *PCSK1*^10^. At the other four genes, heterozygous PTVs conferred higher risk for adult BMI: *BSN* (effect=3.05 kg/m^2^, se=0.54, *P*=2×10^-8^, carrier N=65), *TOX4* (3.61, se=0.71, *P*=3.1×10^-7^, carrier N=39), *APBA1* (2.08, se=0.42, *P*=6.1×10^-7^, carrier N=111) and *ATP13A1* (1.82, se=0.37, *P*=1.1×10^-6^, carrier N=139). For two of these genes, *BSN* and *ATP13A1*, we also found supporting evidence from common genetic variants at the same locus associated with BMI **(Figure S3)** – non-coding alleles ∼200kb upstream of *BSN* (rs9843653, MAF=0.49, beta=-0.13 kg/m^2^, *P*=9.5×10^-46^) and 400kb upstream of *ATP13A1* (rs72999063, MAF=0.16, beta=0.09 kg/m^2^, *P*= 3.2×10^-13^, **Table S2**). Both of these GWAS signals were also associated with blood expression levels of *BSN* and *ATP13A1*, respectively^16^ **(Table S2)**, and the BMI associations were replicated in independent GWAS data from the GIANT consortium^9^ **(Figure S4, Table S2)**. We found no evidence of rare variant associations with BMI for any other genes at these GWAS loci **(Table S3)**.

We aimed to replicate our novel gene-burden rare variant associations in independent WES data from 167,359 individuals of non-European ancestry from the Mexico City Prospective Study (MCPS)^17, 18^ and the Pakistan Genomic Resource (PGR) study **(Table S4, Figure 1)**. We observed supportive evidence for two of the four novel genes identified above – 32 *BSN* PTV carriers had a mean 2.8 kg/m^2^ (se=0.84, *P*=9.4×10^-4^) higher BMI than non-carriers, and 20 *APBA1* PTV carriers had a mean 2.33 kg/m^2^ (se=1.05, *P*=0.03) higher BMI. These effect sizes were remarkably similar to those observed in UK Biobank (3.05 kg/m^2^ and 2.08 kg/m^2^ for *BSN* and *APBA1*, respectively).

**Figure 1.**
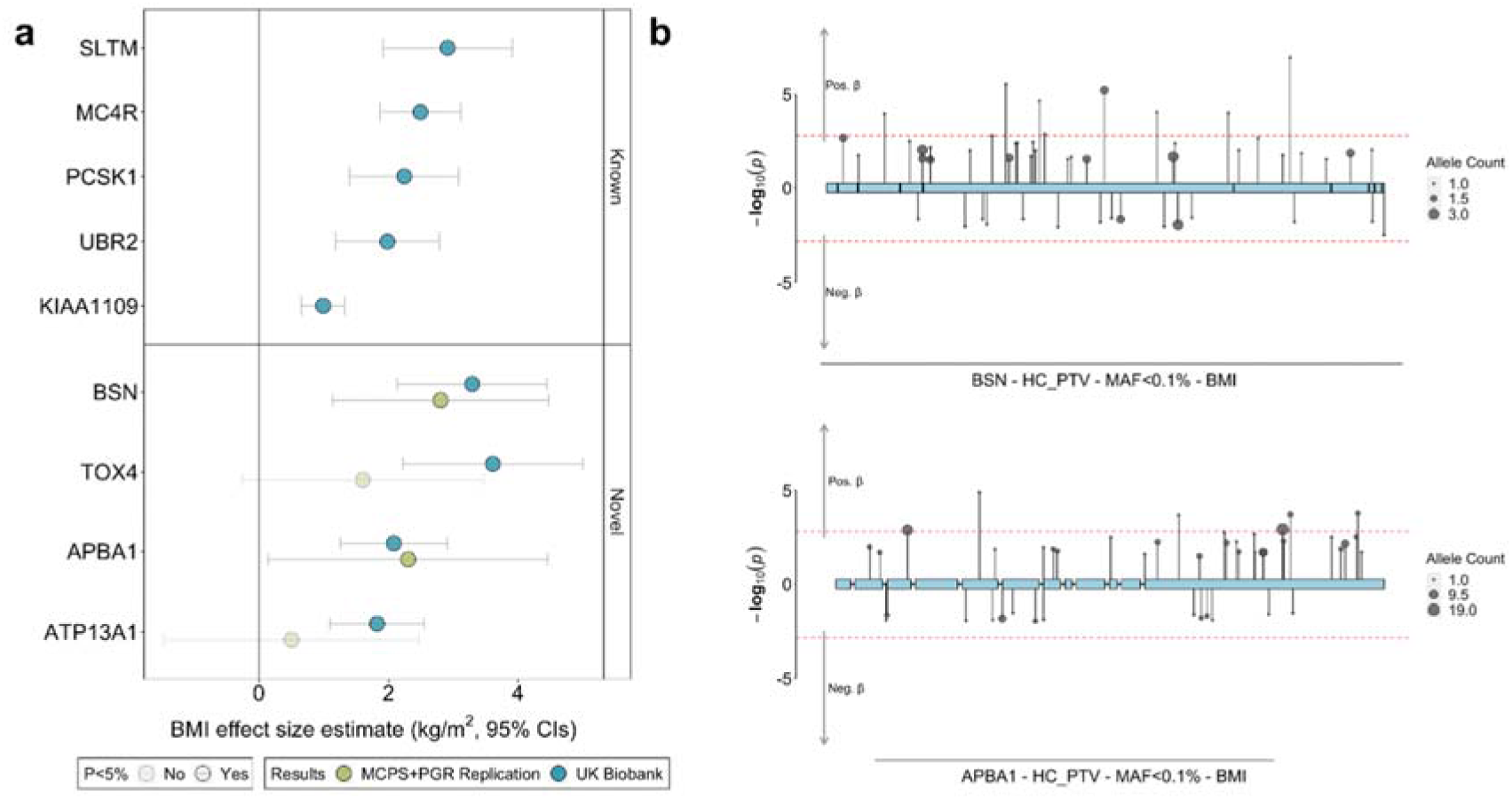
Discovery and replication of novel rare variant associations with BMI in UK Biobank. (a) Effect size estimates have been converted to kg/m^2^. Extended data can be found in Table S1 and S4. (b) Variant-level associations between HC PTVs in *BSN* and *APBA1* and BMI. The Y-axis shows trait increasing effects with a -1*log (10) p-value and trait decreasing effects with a log(10) p-value. Dashed line denotes variants reaching a nominal significance threshold P<0.05.

The effect of *BSN* is larger than any previously reported ExWAS gene on BMI **(Figure 2)**, and substantially increased the risks of obesity in UK Biobank; (*BSN* OR=3.04 [95% confidence interval 1.87-4.94], *P*=7.7×10^-6^, 49% case prevalence; *APBA1* OR=2.14 [1.46-3.13], *P*=8.5×10^-5^, 41% case prevalence) and for *BSN* also increased the risk of severe obesity (OR=6.61 [3.01-14.55], *P*=2.6 ×10^-6^, 11% case prevalence) but not for *APBA1* (OR=1.91 [0.70-5.19], *P*=0.20, 4% case prevalence, **Figure 3**). Association statistics for individual variants in *BSN* and *APBA1* in UK Biobank are shown in **Figure 1B** and **Table S5**. The gene-level associations between *BSN* and *APBA1* and BMI were not driven by single high-confidence (HC) PTVs **(Table S6)**, and carriers appeared to be geographically dispersed across the UK **(Figure S5)**.

**Figure 2.**
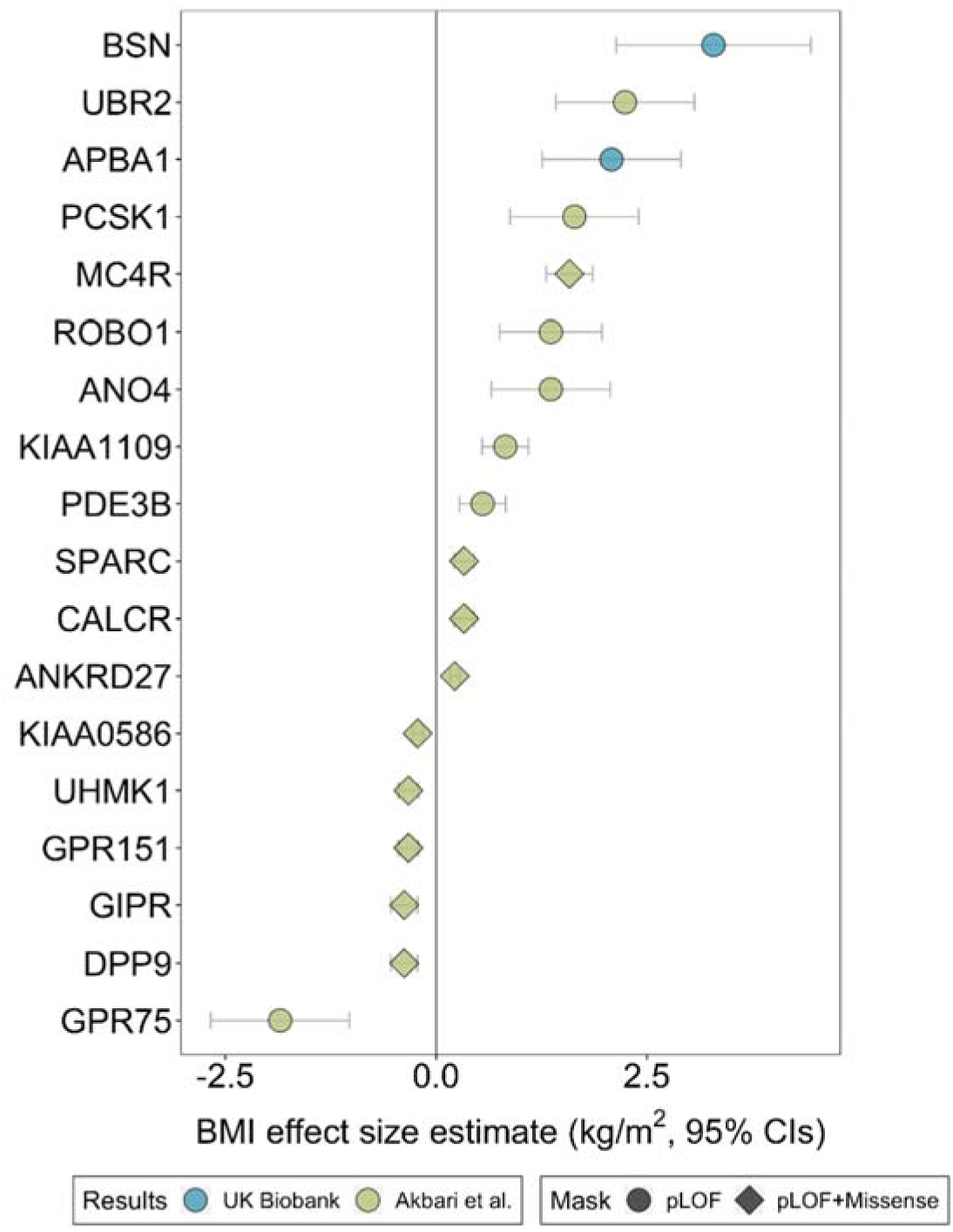
Comparison of effect between replicated associations and previously reported associations^10^. The BMI effect size estimates are based on UK Biobank participants only.

**Figure 3.**
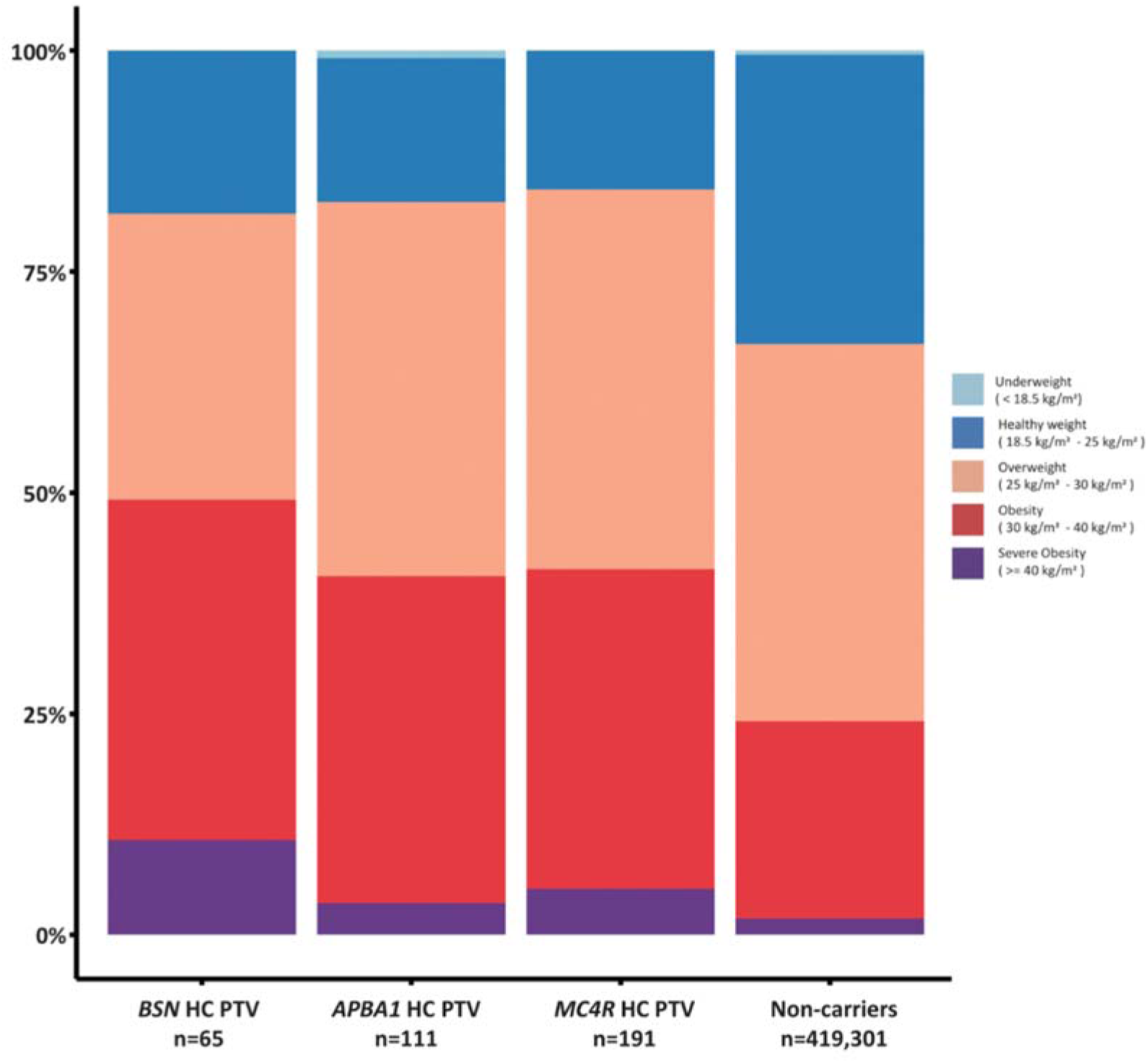
Distribution in BMI categories for the carriers and non-carriers of *BSN*, *APBA1* or *MC4R* HC PTV carriers. The BMI categories are classified according to the WHO’s guidance.

We next sought to understand the broader phenotypic profile of carriers of PTVs in *BSN* and *APBA1*. Both genes showed diverse associations with body composition, with higher fat and lean mass across body compartments **(Table S7)**, but no association with adult height (*P*>0.05) or waist-to-hip ratio adjusted for BMI (*P*>0.05). In contrast to almost all previously reported obesity-associated genes, neither *BSN* or *APBA1* exhibited any association with childhood body size or puberty timing (*P*>0.05), suggesting adult-onset effects on body weight based on the phenotypes available in UK Biobank. In UK Biobank, carriers of PTVs in *BSN* also had higher risk of type 2 diabetes (T2D) – *BSN* OR=3.03 (95% CI [1.60-5.76], *P*= 7.1×10^-4^, 18% case prevalence) – an effect size comparable to previously reported genes for T2D^19, 20^. A broader phenome-wide analysis across 11,693 traits revealed a number of other associations **(Table S8)**; notably *BSN* PTV carriers had a substantially higher risk of non-alcoholic fatty liver disease (NAFLD) - as defined by a Fatty Liver Index (FLI) ≥ 60^21^ or Hepatic Steatosis Index (HSI) > 36^22^ – compared to non-carriers (OR=3.73 (95% CI [2.26-6.16], *P*=8.4×10^-7^, 45% case prevalence).

Previous studies reported that common BMI-associated alleles did not modify the penetrance of rare variants on BMI or obesity^10^. To evaluate whether this was true also for *BSN* and *APBA1*, we created a common variant polygenic score (PGS) in UK Biobank, using individual variant effect estimates obtained from independent GIANT consortium GWAS data^9^. By testing the interaction between the PGS and rare variant carrier status in a linear regression model, we observed significant effect modification by *BSN* PTVs (*P*= 0.01, **Figure S6**), but not *APBA1* (*P*=0.22). Carriers of *BSN* PTVs showed double the effect size of the PGS on BMI (0.6 BMI standard deviations per unit increase in PGS, equivalent to 2.9 kg/m^2^) than non-carriers (0.3 standard deviations, equivalent to 1.4 kg/m^2^).

To explore the putative biological mechanisms through which *BSN* and *APBA1* might exert their effects, we first characterized the plasma proteomic signature of PTV carriers using Olink data on 1,463 circulating proteins available in ∼50,000 UK Biobank participants^23, 24^. We identified 6 and 17 PTV carriers with available proteomics data for *BSN* and *APBA1*, respectively. No plasma proteins were associated with *APBA1* carrier status after multiple-test correction (*P*<3.42 ×10^-5^ (0.05/1,463)), however *BSN* PTV carriers had higher levels of lymphotoxin alpha (LT-α, previously known as TNF-β) than non-carriers (effect=1.07, se=0.183, *P*=5.3×10^-9^) **(Table S9)**. Furthermore, circulating LT-α levels were positively associated with BMI (1.18 kg/m^2^ per 1 SD higher LT-α, *P*=7.6×10^-1^^22^) and common genetic variants at the *LTA* locus were associated with BMI (rs3130048, MAF=0.72, beta=-0.10 kg/m^2^/allele, *P*=1.10×10^-23^). We repeated these analyses using the common BMI-associated variant (rs9843653) at *BSN* and identified 23 associated proteins, the most significant of which was Semaphorin-3F (-0.03 SD per BMI-increasing allele, *P*=6.7×10^-45^), a member of the semaphorin family which has been previously implicated in obesity etiology^25^. In total, 10 of these 24 protein-encoded genes (including *SEMA3F* and *LTA*) were also implicated by common variant signals for BMI (**Table S10**).

Finally, we explored the functional consequences of *BSN* deletion, which is highly expressed in the brain, by generating CRISPR-Cas9 edited human iPSC hypothalamic neurons that were heterozygous for *BSN* loss of function **(Methods)**. Numbers of differentiated *BSN*^+/-^ cells were lower than WT cells (2,924 *BSN*^+/-^ and 18,010 WT); however, on visual inspection there was no apparent morphological effect on neuronal differentiation **(Figure S7)**. To assess transcriptional differences between these cell populations, we performed single nucleus RNAseq across all 20,934 hypothalamic cells differentiated from human iPSCs. A UMAP plot of the cells identified eight distinct clusters **(Figure S8)**, of which two clusters (clusters 1 and 5, encompassing 4,991 cells) were enriched for *RBFOX3* (*NeuN*) expression, a marker for mature neurons. These two clusters were also enriched for expression of *BSN* and its binding partner *Piccolo* (*PCLO*), and were therefore separated and re-clustered for further analysis **(Figure S8)**. This produced a further five clusters, including 2,712 WT and 343 *BSN*^+/-^ cells, which we selected for analysis of differential gene expression (defined by corrected *P*<0.05 and Log2FC > 1 or < -1). These analyses highlighted 251 differentially-expressed genes across one or more of the five clusters (**Table S11**), including genes with established roles in obesity regulation, such as members of the semaphorin gene family^25^ and *ALK*^26^. Pathway enrichment analyses across differentially-expressed genes highlighted a number of biological processes **(Table S12)**, with ‘synapse organization’ and ‘negative regulation of neuron projection development’ the most significantly enriched pathways in clusters 1 and 2, respectively. Collectively, differentially-expressed genes within these two clusters (1 & 2) were also enriched for common variant associations with BMI **(Table S13-14)**.

## Discussion

We identified that rare PTVs in *APBA1* and *BSN* are associated with a substantial increase in adult BMI and higher risk of obesity in adults, but not in childhood. Rare PTVs in *BSN* were also found to be associated with higher risks for T2D and NAFLD. The associations with adult BMI were confirmed in independent cohorts and also supported by mapping of common variant signal to whole-blood eQTLs for *APBA1* and *BSN*.

*APBA1* and *BSN* are among the few genetic determinants of adult-onset obesity. Although childhood adiposity was assessed here by subjective recall, this trait is reported to show high genetic correlation with measured childhood BMI and hence is a valid indicator for genetic analyses^27^. *APBA1* encodes a neuronal adapter protein that interacts with the Alzheimer’s disease-associated gene amyloid precursor protein (*APP*). It has a putative role in signal transduction as a vesicular trafficking protein with the potential to couple synaptic vesicle exocytosis to neuronal cell adhesion^28^. *BSN* encodes Bassoon, a scaffolding protein essential for the organization of the presynaptic cytoskeleton and for exocytosis-mediated neurotransmitter release^29^. *Bsn* knockout in mice reduces excitatory synaptic transmission because vesicles are unable to efficiently fuse with the synaptic membrane^30^. *BSN* is expressed primarily in the brain and is reportedly upregulated in frontal lobes of patients with multiple system atrophy, a progressive neurodegenerative disease^31^. Furthermore, rare predicted-damaging missense mutations in *BSN* were reported in four patients with progressive supranuclear palsy-like syndrome with features of multiple system atrophy and Alzheimer’s disease^32^. Hence, the links we identify here with adult-onset (rather than childhood-onset) obesity may be consistent with the putative roles of *APBA1* and *BSN* in ageing-related neurosecretory vesicle dysfunction and neurodegenerative disorders.

Previous studies reported additive effects of common and rare susceptibility alleles on BMI^10^, but no evidence for epistatic interactions that are indicative of biological interaction. Notably, we found that carriers of rare PTVs in *BSN* showed enhanced susceptibility to the influence of a common genetic polygenic risk score for adult BMI. The mechanistic basis for this statistical interaction is unclear. However, as the common genetic susceptibility to obesity is thought to act predominantly via the central regulation of food intake^9, 33^, we hypothesize that *BSN* might have widespread involvement in the synaptic secretion of neurotransmitters that suppress appetite and increase satiety.

In conclusion, rare genetic disruption of *APBA1* and *BSN* have larger impacts on adult BMI and obesity risk than heterozygous disruption of any other described obesity risk gene. Rare PTVs in *APBA1* and *BSN* appear to preferentially confer risk to adult-onset obesity, which we propose might be due to widespread vesicular dysfunction leading to reduced synaptic secretion of neurotransmitters that suppress food intake.

## Supporting information

Supplementary Informations

## Data Availability

The UK Biobank phenotype and whole-exome sequencing data described here are publicly available to registered researchers through the UKB data access protocol. Information about registration for access to the data is available at: https://www.ukbiobank.ac.uk/enable-your-research/apply-for-access. Data for this study were obtained under Resource Application Numbers 26041 and 9905.
The Mexico City Prospective Study welcomes open access and collaboration data requests from bona fide researchers. For more details on accessibility, the study's Data and Sample Sharing policy may be downloaded (in English or Spanish) from https://www.ctsu.ox.ac.uk/research/mcps. Available study data can be examined in detail through the study's Data Showcase, available at https://datashare.ndph.ox.ac.uk/mexico/.

## Acknowledgements

**Competing interests:** Z. F.-H., Q.W., K.R.S, D.S.P., and S.P. are current employees and/or stockholders of AstraZeneca. J.R.B.P and E.G are employees and shareholders of Adrestia Therapeutics.

**Funding:** The MCPS has received funding from the Mexican Health Ministry, the National Council of Science and Technology for Mexico, the Wellcome Trust (058299/Z/99), Cancer Research UK, British Heart Foundation, and the UK Medical Research Council (MC_UU_00017/2). These funding sources had no role in the design, conduct, or analysis of the study or the decision to submit the manuscript for publication.

We thank the participants and investigators in the UKB study who made this work possible (Resource Application Number 26041; 9905) the UKB Exome Sequencing Consortium (UKB-ESC) members AbbVie, Alnylam Pharmaceuticals, AstraZeneca, Biogen, Bristol-Myers Squibb, Pfizer, Regeneron and Takeda for funding the generation of the data; the Regeneron Genetics Center for completing the sequencing and initial quality control of the exome sequencing data; and the AstraZeneca Centre for Genomics Research Analytics and Informatics team for processing and analysis of sequencing and phenotype data.

## Data Accessibility

The UK Biobank phenotype and whole-exome sequencing data described here are publicly available to registered researchers through the UKB data access protocol. Information about registration for access to the data is available at: https://www.ukbiobank.ac.uk/enable-your-research/apply-for-access. Data for this study were obtained under Resource Application Numbers 26041 and 9905.

The Mexico City Prospective Study welcomes open access and collaboration data requests from bona fide researchers. For more details on accessibility, the study’s Data and Sample Sharing policy may be downloaded (in English or Spanish) from https://www.ctsu.ox.ac.uk/research/mcps. Available study data can be examined in detail through the study’s Data Showcase, available at https://datashare.ndph.ox.ac.uk/mexico/.

## Methods

### UK biobank data processing and quality control

We employed the same processing strategies as outlined in our previous paper to analyse the whole-exome sequencing data and perform quality control steps^19^. We queried whole-exome sequencing data from 454,787 individuals in the UK Biobank^34^, excluding those with excess heterozygosity, autosomal variant missingness on genotyping arrays >= 5%, or those not included in the subset of phased samples as defined by Bycroft et al^13^.

The whole-exome sequencing data was stored as population-level VCF files, aligned to GRCh38, and accessed through the UKBB RAP. In addition to the quality control measures already applied to the released data, which were described by Backman et al.^34^, we conducted several extra QC procedures. Firstly, we used ‘bcftools norm’ ^35^ to split the multiallelic sites and left-correct and normalise InDels. Next, we filtered out variants that failed our QC criteria, including: 1) read depth < 7, 2) genotype quality < 20, and 3) binomial test p-value for alternate allele reads versus reference allele reads <= 0.001 for heterozygous genotypes. For InDel genotypes, we only kept variants with read depth >= 10 and genotype quality >= 20. Variants that failed QC criteria were marked as missing (i.e.,./.). After filtering, variants where more than 50% of genotypes were missing were excluded from downstream analyses^19^.

The remaining variants underwent annotation using ENSEMBL Variant Effect Predictor (VEP) v104^36^ with the ‘-everything’ flag, and additional plugins for REVEL^14^, CADD^37^, and LOFTEE^38^. For each variant, a single ENSEMBL transcript was prioritised based on whether the annotated transcript was protein-coding, MANE select v0.97^39^, or the VEP Canonical transcript. The individual consequence for each variant was then prioritised based on severity as defined by VEP. Stop-gained, splice acceptor, and splice donor variants were merged into a combined Protein Truncating Variant category, while annotations for missense and synonymous variants were adopted directly from VEP. We only included variants on autosomes and the X chromosome that were within ENSEMBL protein-coding transcripts and within transcripts included on the UKBB WES assay in our downstream analysis.

Our analyses focused primarily on individuals of European genetic ancestry, and we excluded those who withdrew consent from the study, resulting in a final cohort of 419,668 individuals.

### Exome-wide gene burden testing in the UK Biobank

We used BOLT-LMM v2.3.6^15^ as our primary analytical tool to conduct the gene burden test. To run BOLT-LMM, we first queried a set of genotypes with MAC > 100 which derived from the genotyping arrays for the individuals with the WES data to build the null model. To accommodate BOLT-LMM’s requirement for imputed genotyping data rather than per-gene carrier status, we developed dummy genotype files where each gene was represented by a single variant. We then coded individuals with a qualifying variant within a gene as heterozygous, regardless of the total number of variants they carried in that gene. We then created the dummy genotypes for the MAF < 0.1% high confidence PTVs as defined by LOFTEE, missense variants with REVEL > 0.5 and missense variants with REVEL > 0.7. We then used BOLT-LMM to analyse phenotypes, using default parameters except for the inclusion of the ’lmmInfOnly’ flag. In addition to the dummy genotypes, we also included all individual markers contained in WES data to generate the association test statistics for individual variants. We used age, age^2^, sex, the first 10 principal components as calculated by Bycroft et al.^13^, and the WES released batch (50k, 200k, 450k) as covariates.

To check whether there is a single variant driving the association, we performed a leave-one-out analysis for *BSN* and *APBA1* using linear regression in R v3.6.3 by dropping the HC PTV variants contained in our analysis one by one. In addition, we also checked the geographic distribution of *APBA1* and *BSN* HC PTV carriers.

### Replication of the findings in two independent non-European cohorts

We tried to replicate our findings for the four novel genes in two independent non-European exome-sequenced cohorts: Mexico City Prospective Study (MCPS) and The Pakistan Genomic Resource (PGR) study.

Mexico City Prospective Study is a cohort study of 159,755 adults of predominantly Admixed American ancestry. Participants were recruited between 1998 and 2004 aged 35 years or older from two adjacent urban districts of Mexico City. Phenotypic data were recorded during household visits, including height, weight, and waist and hip circumferences. Disease history was self-reported at baseline, and participants are linked to Mexican national mortality records. The cohort has been described in detail elsewhere^17, 18^. The MCPS study was approved by the Mexican Ministry of Health, the Mexican National Council for Science and Technology, and the University of Oxford.

The Pakistan Genomic Resource study has been recruiting participants aged 15-100 years as cases or controls via clinical audits for specific conditions since 2005 from over 40 centres around Pakistan. Participants were recruited from clinics treating patients with cardiometabolic, inflammatory, respiratory, or ophthalmological conditions. Information on lifestyle habits, medical and medication history, family history of diseases, exposure to smoking and tobacco consumption, physical activity, dietary habits, anthropometry, basic blood biochemistry and ECG traits were recorded during clinic visits. DNA, serum, plasma, and whole-blood samples were also collected from all study participants. The institutional review board at the Center for Non-Communicable Diseases (IRB: 00007048, IORG0005843, FWAS00014490) approved the study and all participants gave informed consent.

Exome sequencing data for 141,046 MCPS and 37,800 PGR participants were generated at the Regeneron Genetics Center and passed Regeneron’s initial quality control (QC) that included identifying sex discordance, contamination, unresolved duplicate sequences, and, for MCPS, discordance with microarray genotype data. Genomic DNA underwent paired-end 75–base pair whole-exome sequencing at Regeneron Pharmaceuticals using the IDT xGen v1 capture kit on the NovaSeq6000 platform. Conversion of sequencing data in BCL format to FASTQ format and the assignments of paired-end sequence reads to samples were based on 10-base barcodes, using bcl2fastq v2.19.0.

These exome sequences were processed at AstraZeneca from their unaligned FASTQ state. A custom-built Amazon Web Services cloud computing platform running Illumina DRAGEN Bio-IT Platform Germline Pipeline v3.0.7 was used to align the reads to the GRCh38 genome reference and perform single-nucleotide variant (SNV) and insertion and deletion (indel) calling. SNVs and indels were annotated using SnpEff v4.3^40^ against Ensembl Build 38.92. All variants were additionally annotated with their gnomAD MAFs (gnomAD v2.1.1 mapped to GRCh38)^38^.

To further QC the sequence data, all MCPS and PGR exomes underwent a second screen using AstraZeneca’s bioinformatics pipeline which has been described in detail previously^41^. Briefly, we excluded from analysis sequences that achieved a VerifyBamID freemix (contamination) level of more than 4%, where inferred karyotypic sex did not match self-reported gender, or where less than 94.5% of the consensus coding sequence (CCDS release 22) achieved a minimum tenfold read depth. We further removed one individual from every pair of genetic duplicates or monozygotic twins with a kinship coefficient > 0.45.

Kinship coefficients were estimated from exome genotypes using the --kinship function from KING v2.2.3^42^. For MCPS we additionally excluded sequences with an average CCDS read depth at least two standard deviations (SD) below the mean. After the above quality control steps, there remained 139,603 (99.0%) MCPS and 37,727 (99.3%) PGR exomes.

In MCPS, we predicted the genetic ancestry of participants using PEDDY v0.4.2^43^, with 1000 Genomes Project sequences as population references^44^ and retain individuals with a predicted probability of Admixed American ancestry ≥ 0.95 who were within 4 SD of the means for the top four principal components (PCs). In PGR we retained individuals with a predicted probability of South Asian ancestry ≥ 0.95 who were within 4 SD of the means for the top four PCs. Following ancestry filtering, 137,059 (97.2%) MCPS and 36,280 (95.5%) PGR exomes remained.

We assessed the association between BMI and weight quantitative traits with genotypes at the four proposed novel genes of interest using a previously described gene-level collapsing analysis framework, implementing a protein truncating variant (PTV) collapsing analysis model^41^. We classified variants as PTVs if they had been annotated by SnpEff as: exon_loss_variant, frameshift_variant, start_lost, stop_gained, stop_lost, splice_acceptor_variant, splice_donor_variant, gene_fusion, bidirectional_gene_fusion, rare_amino_acid_variant, and transcript_ablation.

We applied minor allele frequency filters to target rare variants; MAF < 0.001 in gnomAD (overall and every population except OTH) and a leave-one-out MAF < 0.001 among our combined case and control test cohort. For variants to qualify they had to also meet the following QC filters: minimum site coverage 10X; annotation in CCDS transcripts (release 22); at least 80% alternate reads in homozygous genotypes; percent of alternate reads in heterozygous variants ≥ 0.25 and ≤ 0.8; binomial test of alternate allele proportion departure from 50% in heterozygous state P > 1 × 10^−6^; GQ ≥ 20; FS ≤ 200 (indels) ≤ 60 (SNVs); MQ ≥ 40; QUAL ≥ 30; read position rank sum score ≥ −2; MQRS ≥ −8; DRAGEN variant status = PASS and <0.5% test cohort carrier QC failure. If the variant was observed in gnomAD exomes, we also applied the filters: variant site achieved tenfold coverage in ≥ 25% of gnomAD exomes; achieved exome z-score ≥ −2.0; exome MQ ≥ 30 and random forest probability that the given variant is a true SNV or indel > 0.02 and > 0.01 respectively^45^.

For the quantitative traits, and for each gene, the difference in mean between the carriers and noncarriers of PTVs was determined by fitting a linear regression model, correcting for age and sex. In addition to calculating individual statistics for MCPS and PGR, we also meta-analysed the individual study effect sizes to generate a combined replication statistic by using an inverse-variance fixed effect meta-analysis implemented in R using the rma.uni() function from the package ‘metafor’ v3,8-1^46^

### Phenome-wide analysis in UKBB

We included binary and quantitative traits made available in the June 2022 UKB data release, harmonizing the phenotype data as previously described^41^. This resulted in 11,690 phenotypes for analysis; as available on https://azphewas.com. Based on clinical relevance we derived an additional three phenotypes.

For the purposes of UKB phenome-wide analyses of the four putatively novel genes, the same data generation and QC processes described for MCPS and PGR were applied to UKB exomes. Following Regeneron and AstraZeneca QC steps, 445,570 UKB exomes remained. The phenome-wide analysis was performed in UKB participants of predominately EUR descent, whom we identified based on a PEDDY-derived predicted probability of European ancestry ≥ 0.95 who were within 4 SD of the means for the top four PCs. Based on the predicted ancestry pruning, 419,391 UKB exomes were included in the phenome-wide analyses of the four priority genes.

As described previously, we assessed the association between the 11,693 phenotypes with genotypes at the four genes of interest, again using a PTV collapsing analysis model^41^, classifying variants as PTVs using the same SnpEff definitions as described for the MCPS and PGR analysis. For variants to qualify for inclusion in the model, we applied the same MAF and QC filters used in MCPS and PGR, with the exception that due to the larger sample size of UKB, only <0.01% of the test cohort carriers were permitted to fail QC.

### Association testing for other anthropometry phenotypes and protein expression level

We ran association tests between *APBA1* and *BSN* HC PTV carriers and BMI-associated common variant (rs9843653) at the *BSN* locus carriers and a list of anthropometry phenotypes in R v3.6.3 **(Table S5)** including the same covariates we used in our exome-wide gene burden tests. We acquired the normalised protein expression data generated by the Olink platform from the UKBB RAP^23, 24^. The detailed Olink proteomics assay, data processing and quality control were described by Sun et al. ^23^ For the association tests between *APBA1* and *BSN* PTV carriers and BMI-associated common variant (rs9843653) at the *BSN* locus carriers and 1,463 protein expression levels, we added age2, age*sex, age^2^*sex, Olink batch, UK Biobank centre, UK Biobank genetic array, number of proteins measured and the first 20 genetic principal components (PCs) as covariates as suggested by Sun et al.^23^ We chose the Bonferroni corrected p-value (*P*<3.42 ×10^-5^ (0.05/1,463)) as the threshold for the significant associations.

### BMI GWAS lookup and downstream analyses

Identified genes were queried for proximal BMI GWAS signals, using data from the UK Biobank, if within 500kb up-or downstream of the gene’s start or end site. Any such signals were further replicated in an independent BMI GWAS^9^.

We also performed colocalisation tests, using the Approximate Bayes Factor (ABF) method in the R package “coloc” (version 5.1.0, 8) and blood gene expression level data from the eQTLGen study^16^. Genomic regions were defined as ±500kb around each gene and loci exhibiting an H4 posterior probability >0.5 were considered to show evidence of colocalisation.

Finally, we also used the GWAS data to calculate gene-level common variant associations, using MAGMA^47^. To do this, we used all common but non-synonymous (coding) variants within a given gene. Gene-level scores were further collapsed into pathway-level associations where appropriate.

### Interaction effect between PGS and PTV carrier status

To examine whether there is an interaction effect between the PTV carrier status of *BSN* and *APBA1*, we included an interaction term between PGS and the carrier status of *BSN* and *APBA1* PTVs in a linear regression adjusted for sex, age and age squared and the first 10 PCs.

The polygenic score (PGS) was constructed for 419,581 individuals of white European ancestry who had both genotype and exome sequencing data and a BMI record in the UKBB. We used summary statistics of BMI from Locke et al.^9^, which included samples not in the UKBB. Data was downloaded from the GIANT consortium. The summary statistics included 2,113,400 SNPs with at least 50,0000 samples in a cohort of 322,154 participants of European ancestry. For the genotype data of UKBB participants, a light quality check procedure was applied, where SNPs were removed if they had a MAF <0.1%, HWE <1e-6 or variants with more than 10% missingness genotypes. Additionally, SNPs that were mismatched with those in the summary statistics (same rsID but different chromosome or positions) were excluded. We used lassosum v4.0.5^48^ to construct the PGS. The R squared of the model including the PGS regressed on rank-based inverse normal transformed BMI and adjusted for sex, age and age squared and the first 10 PCs as covariates was 11%.

### Cell lines and routine cell culture

The KOLF2.1J human-induced pluripotent stem cell line and its derivatives^49^ were maintained on Geltrex (Thermo Fisher Scientific A1413202) coated plates in supplemented StemFlex media (Thermo Fisher Scientific A3349401) with daily medium changes. For passaging, the cells were washed with PBS and treated with TrypLE Express (Gibco, 12604021) at 37℃ for 3 min. The cells were re-suspended in StemFlex media supplemented with 10 μM ROCK Inhibitor Y-27632 dihydrochloride (Stemcell Technologies, 72304). ROCK inhibitor was removed the following day with growth medium without the Y-27632. Unless otherwise stated, cells were split at a 1:5 ratio. The absence of mycoplasma was confirmed using an EZ-PCR Mycoplasma Test Kit (Biological Industries, 20-700-20) following the manufacturer’s instructions.

### CRISPR-Cas9-mediated targeting of BSN

Two different small guide RNAs (sgRNA) with high predicted on-target and low predicted off-target activity were designed using CRISPick (https://portals.broadinstitute.org/gppx/crispick/public). For the production of sgRNAs, a 120 nucleotide oligo (Integrated DNA Technologies Inc.) including the SP6 promoter, sgRNA sequences, and scaffold region were used as a template for synthesis by *in vitro* transcription using the MEGAscript SP6 kit (Thermo Fisher, AM1330) as previously described^50^. The resulting sgRNAs were purified using the E.Z.N.A miRNA purification kit (Omega Bio-tek, R7034-01), eluted in RNase-free water, and stored at −80℃. Since sgRNAs vary in their efficacy, the relative cutting efficiencies of the two sgRNAs were tested in *in vitro* cleavage assays as previously described^50^. We selected the sgRNAs that showed activity at the lowest Cas9 concentration for transfection into hPSC cells. Single-stranded oligodeoxynucleotides (ssODN) templates (100bp) were constructed by IDT containing target mutations in the middle and silent mutations within PAM sites. All sequences of the primers, sgRNA, ssODN donors used in the study are listed in **(Table S15)**.

### CRISPR-Cas9 ribonucleoprotein (RNP) complex-mediated editing in hESCs

To genetically edit the KOLF2.1J cells by homology-directed repair (HDR)^51, 52^, 3 μg purified sgRNA was mixed with 4 μg of recombinant Cas9 nuclease (IDT 1081060) for 45 min at room temperature to form stable ribonucleoprotein (RNP) complexes. The complex together with 1μl of 100 μM ssODN was then transferred to a 20μl single-cell suspension of 2 × 10^5^ hESCs in P3 nucleofection solution and electroporated using Amaxa 4D-Nucleofector™ (Lonza) with program CA137. Transfected cells were seeded onto Geltrex-coated 24 well plates containing a pre-warmed StemFlex medium containing Revitacell (100x, Gibco A2644501) and Penicillin/Streptavidin (ThermoFisher Scientific, 15140-122). HDR enhancer (IDT 1081072) was added to the cells at a 30μM final concentration for each well. The following day medium was changed to growth medium without Pen/Strep and Revitacell. To increase HDR efficiency, cells were cultured under cold shock conditions (32°C at 5% CO_2_ in air atmosphere) for 48hr post transfection. Cells were given approximately 5-6 days to recover before single cells were then distributed into multiple Geltrex (1:40)-coated 96 well plates by an Aria-Fusion sorter with a 100 μm nozzle. After ∼2 weeks, viable clonally-derived colonies were consolidated into duplicate 96 well plates to allow parallel cell cryopreservation and genomic DNA extraction as previously described^50, 52^.

### Generation and sequencing of pooled amplicons

Genomic DNA (gDNA) was extracted using HotShot buffer as previously described^50^. The target regions were amplified from gDNA using locus-specific primers to generate amplicons approximately 150-200 bp in length. These “first-round” primers contained universal Fluidigm linker sequences at their 5’-end with the following sequences: Forward primer: 5’-acactgacgacatggttctaca -3’, Reverse primer: 5’-tacggtagcagagacttggtct-3’. Specifically, 20 μl PCR reactions were set up in 96 well plates using 0.5U Phusion Hot Start II High-Fidelity DNA Polymerase (ThermoFisher Scientific, F-549L), 2 μl of extracted gDNA as template, 2 μl 5x GC buffer, 0.2 mM dNTPs, 2μM primers, and 3% DMSO, and run on the following programme: 98°C 30sec, followed by 24 cycles of (95°C 10 sec, 72°C 20 sec/ decreased by 0.5°C per cycle, 72°C 15 sec) than 12 cycles of (98°C 10sec, 60°C 30 sec, 72°C 15 sec) and 72°C 5 min. In the second round of PCR (indexing PCR), Fluidigm barcoding primers were attached to the amplicons to uniquely identify each clone. 2 μl linker PCR product diluted 1:10 was transferred to another 96-well PCR plate to perform this indexing PCR in 10 μl reactions containing 0.8 μM of Fluidigm barcoding primers, 2 μl 10x GC buffer, 0.2 mM dNTPs 3% DMSO and 0.5U Phusion Hot Start II polymerase. The PCR programme was 95°C 2 min, 16 cycles of (95°C 20 sec, 60°C 20 sec, 72°C 25 sec), 72°C 3 min. For sequencing library preparation, barcoded PCR products were combined in equal proportion based on estimation of band intensity on a 2% agarose gel, and the combined pool of PCR products was purified in a single tube using Ampure XP beads (Invitrogen 123.21D) at 1:1 (V/V) to the pooled sample and eluted in 25 μl of water according to the manufacturer’s instructions. Library purity was confirmed by nanodrop, and final library concentration was measured using the Agilent Bioanalyzer (High Sensitivity Kit, Agilent 5067-4626) and diluted to 20 nM. Pooled libraries could be combined with other library pools adjusted to 20 nM, and the resulting “superpool” volume was adjusted to a final volume of 20 μl before sequencing which is performed by the Genomics Core, Cancer Research UK Cambridge Institute. GenEditID platform^52^ was used for identification of the BSN P399X heterozygous and wild type (WT) clones.

### Hypothalamic neuron differentiation protocol

Gene edited *BSN* P399X heterozygous and WT clones were differentiated into hypothalamic-like neurons as previously described^53, 54^. Briefly, cells were cultured overnight on 10cm plate Geltrex coated plates (9.5 ×10^5^ cells/well for 6-well plates) in Stemflex^TM^ with 10 μM ROCK inhibitor. Next day, neuroectoderm differentiation was initiated by dual SMAD inhibition using XAV939 (Stemgent 04-1946), LDN 193289 (Stemgent 04-0074) and SB 431542 (Sigma Aldrich S4317) and Wnt signaling inhibition using XAV939 (Stemgent 04-1946) in an in-house neural differentiation N2B27 medium^54^. From day 2 to day 7, cells were directed ‘towards ventral diencephalon’ with Sonic hedgehog activation, by the addition of Smoothened agonist SAG (1μM Thermo Fisher Scientific 56-666) and purmorphamine (PMC, 1 μM Thermo Fisher Scientific 54-022), with SMAD and Wnt inhibition molecules gradually replaced with N2 B27 medium changed every 2 days. At Day 8, the cells were switched into N2B27 with 5 μM DAPT (Sigma Aldrich D5942) to exit cell cycle. On Day 14, the cells were harvested with TrypLE^TM^ supplemented with papain (Worthington LK003176) and re-plated onto laminin–coated 6-well plates at a density of 3×10^6^ cells per well in the presence of maturation medium containing brain-derived neurotrophic factor BDNF (10ng/ml, Sigma) containing N2B27. On day 16 media was changed to Synaptojuice 1 (N2B27, 10ng/ml BDNF, 2 μM PD0332991 (Sigma Aldrich, PZ0199), 5 μM DAPT, 370 μM CaCl_2_ (Sigma Aldrich, 21115), 1 μM LM22A4 (Tocris, 4607), 2 μM CHIR99021 (Cell Guidance Systems, SM13), 300 μM GABA (Tocris, 0344), 10 μM NKH447 (Sigma Aldrich, N3290)). Cells were maintained in Synaptojuice 1 for a week before being changed to Synaptojuice 2 (N2B27, 10ng/ml BDNF, 2 μM, 370 μM CaCl_2_ 1 μM LM22A4, 2 μM CHIR99021). Cells were then maintained in Synaptojuice 2 until day 36, with media renewal every second day throughout the differentiation and maturation period.

### Single nucleus RNA-sequencing

On day 36, cells were dissociated using TrypLE™ and papain mixture, pelleted, and nuclei were isolated following a 10x Genomics standardised protocol for single nucleus RNA Sequencing (NucSeq) as previously reported^55^. Sequencing libraries for the 6 (3 x wild type and 3 x *BSN* P399X Het) single-nuclei suspension samples were generated using 10X Genomics Chromium Single-Cell 3′ Reagent kits (Pleasanton, CA, USA, version 3) according to the standardised protocol. Briefly, nuclear suspensions were loaded onto the chromium chip along with gel beads, partitioning oil, and master mix to generate GEMs containing free RNA. RNA from lysed nuclei was reverse transcribed and cDNA was PCR amplified for 19 cycles. The amplified cDNA was used to generate a barcoded 3′ library according to the manufacturer’s protocol, and paired-end sequencing was performed using an Illumina NovaSeq 6000 (San Diego, CA, USA, read 1: 28 bp and read 2: 91 bp). Library preparation and sequencing was performed by the Genomics Core, Cancer Research UK Cambridge Institute.

### Single-cell clustering and differential gene expression analysis

For the 10X generated NucSeq datasets, Cellranger Version 6.0.1 was used to map sequence reads to the human genome GRCh38 and perform the UMI and gene-level counts against Ensembl gene model V100. The raw count matrices generated by the software were then used for downstream analyses. A downstream analysis on the raw count matrices was performed using the Seurat package version 4.0.3.^56^. Nuclei expressing less than 500 features, or less than 800 transcripts were removed as low-quality reads. Nuclei with more than 10000 different features were removed as these were likely doublets. Any nuclei expressing more than 5% mitochondrial RNA were excluded from the analysis. The SCTransform package was used for normalization and variance stabilization of the data, using regularized negative binomial regression^57^. The data was integrated prior to PCA, followed by unsupervised clustering analysis using the Louvain algorithm and Uniform Manifold Approximation and Projection (UMAP) dimension reduction. Marker genes for each cluster were identified using Wilcoxon’s rank-sum test and receiver-operating curve (ROC) analyses. Adjustment of p-values was performed using Bonferroni correction based on the total number of genes in the dataset. Clusters with a high expression of a conventional neuronal marker *RBFOX3* were separated into a new object and PCA, followed by unsupervised clustering analysis using the Louvain algorithm and Uniform Manifold Approximation and Projection (UMAP) dimension reduction was repeated. After that differential expression of genes between nuclei from the wild type and *BSN* P399X Het nuclei within each cluster was analysed using Negative Binomial GLM fitting and Wald statistics with the help of the DESeq2 package^x258^. The p-values attained by the Wald test were corrected for multiple testing using the Benjamini and Hochberg method to generate adjusted p-values. Genes with a Log2FC of <-1 or >1 and with a p-value adjusted < 0.05 were selected for performing pathway analysis. The Metascape^59^ pathway analysis was used to identify pathways that were either upregulated or downregulated between the wild type and BSN P399X Het nuclei.

## Notes

### Author Declarations

UK Biobank has approval from the North West Multi-centre Research Ethics Committee (REC reference 13/NW/0157, https://www.ukbiobank.ac.uk/media/lcvbdoik/21-nw-0157-favourable-opinion-with-conditions-18-06-2021.pdf) as a Research Tissue Bank (RTB) approval and informed consent (https://www.ukbiobank.ac.uk/media/t22hbo35/consent-form.pdf) was provided by each participant. The MCPS study was approved by the Mexican Ministry of Health, the Mexican National Council for Science and Technology, and the University of Oxford.The institutional review board at the Center for Non-Communicable Diseases (IRB: 00007048, IORG0005843, FWAS00014490) approved the Pakistan Genomic Resource study and all participants gave informed consent.

## References

1. Blüher, M. Obesity: global epidemiology and pathogenesis. Nature Reviews Endocrinology 2019 15:5 15, 288–298 (2019).

2. Health Effects of Overweight and Obesity in 195 Countries over 25 Years. New England Journal of Medicine 377, 13–27 (2017).

3. Di Cesare, M. et al. The epidemiological burden of obesity in childhood: A worldwide epidemic requiring urgent action. BMC Med 17, 1–20 (2019).

4. Zhang, Y. et al. Positional cloning of the mouse obese gene and its human homologue. Nature 372, 425–432 (1994).

5. Loos, R. J. F. & Yeo, G. S. H. The genetics of obesity: from discovery to biology. Nat Rev Genet 23, 120–133 (2022).

6. Vaisse, C., Clement, K., Guy-Grand, B. & Froguel, P. A frameshift mutation in human MC4R is associated with a dominant form of obesity. Nature Genetics 1998 20:2 20, 113–114 (1998).

7. Yeo, G. S. H. et al. A frameshift mutation in MC4R associated with dominantly inherited human obesity. Nature Genetics 1998 20:2 20, 111–112 (1998).

8. Yengo, L. et al. Meta-analysis of genome-wide association studies for height and body mass index in ∼700000 individuals of European ancestry. Hum Mol Genet 27, 3641–3649 (2018).

9. Locke, A. E. et al. Genetic studies of body mass index yield new insights for obesity biology. Nature 2015 518:7538 518, 197–206 (2015).

10. Akbari, P. et al. Sequencing of 640,000 exomes identifies GPR75 variants associated with protection from obesity. Science (1979) 373, (2021).

11. Povysil, G. et al. Rare-variant collapsing analyses for complex traits: guidelines and applications. Nature Reviews Genetics 2019 20:12 20, 747–759 (2019).

12. Stankovic, S. et al. Genetic susceptibility to earlier ovarian ageing increases de novo mutation rate in offspring. medRxiv 2022.06.23.22276698 (2022) doi:10.1101/2022.06.23.22276698.

13. Bycroft, C. et al. The UK Biobank resource with deep phenotyping and genomic data. Nature 2018 562:7726 562, 203–209 (2018).

14. Ioannidis, N. M. et al. REVEL: An Ensemble Method for Predicting the Pathogenicity of Rare Missense Variants. Am J Hum Genet 99, 877–885 (2016).

15. Loh, P. R. et al. Efficient Bayesian mixed-model analysis increases association power in large cohorts. Nature Genetics 2015 47:3 47, 284–290 (2015).

16. Võsa, U. et al. Large-scale cis-and trans-eQTL analyses identify thousands of genetic loci and polygenic scores that regulate blood gene expression. Nature Genetics 2021 53:9 53, 1300–1310 (2021).

17. Tapia-Conyer, R. et al. Cohort Profile: The Mexico City Prospective Study. Int J Epidemiol 35, 243–249 (2006).

18. Ziyatdinov, A. et al. Genotyping, sequencing and analysis of 140,000 adults from the Mexico City Prospective Study. bioRxiv 2022.06.26.495014 (2022) doi:10.1101/2022.06.26.495014.

19. Gardner, E. J. et al. Damaging missense variants in IGF1R implicate a role for IGF-1 resistance in the etiology of type 2 diabetes. Cell Genomics 2, 100208 (2022).

20. Zhao, Y. et al. GIGYF1 loss of function is associated with clonal mosaicism and adverse metabolic health. Nature Communications 2021 12:1 12, 1–6 (2021).

21. Bedogni, G. et al. The fatty liver index: A simple and accurate predictor of hepatic steatosis in the general population. BMC Gastroenterol 6, 1–7 (2006).

22. Lee, J. H. et al. Hepatic steatosis index: A simple screening tool reflecting nonalcoholic fatty liver disease. Digestive and Liver Disease 42, 503–508 (2010).

23. Sun, B. B. et al. Genetic regulation of the human plasma proteome in 54,306 UK Biobank participants. bioRxiv 2022.06.17.496443 (2022) doi:10.1101/2022.06.17.496443.

24. Dhindsa, R. S., et al. Influences of rare protein-coding genetic variants on the human plasma proteome in 50,829 UK Biobank participants. bioRxiv 2022.10.09.511476 (2022) doi:10.1101/2022.10.09.511476.

25. van der Klaauw, A. A. et al. Human Semaphorin 3 Variants Link Melanocortin Circuit Development and Energy Balance. Cell 176, 729–742.e18 (2019).

26. Orthofer, M. et al. Identification of ALK in Thinness. Cell 181, 1246–1262.e22 (2020).

27. Richardson, T. G., Sanderson, E., Elsworth, B., Tilling, K. & Smith, G. D. Use of genetic variation to separate the effects of early and later life adiposity on disease risk: mendelian randomisation study. BMJ 369, (2020).

28. Butz, S., Okamoto, M. & Südhof, T. C. A tripartite protein complex with the potential to couple synaptic vesicle exocytosis to cell adhesion in brain. Cell 94, 773–782 (1998).

29. Tom Dieck, S., et al. Bassoon, a Novel Zinc-finger CAG/Glutamine-repeat Protein Selectively Localized at the Active Zone of Presynaptic Nerve Terminals. Journal of Cell Biology 142, 499–509 (1998).

30. Altrock, W. D. et al. Functional inactivation of a fraction of excitatory synapses in mice deficient for the active zone protein bassoon. Neuron 37, 787–800 (2003).

31. Hashida, H. et al. Cloning and Mapping of ZNF231, a Novel Brain-Specific Gene Encoding Neuronal Double Zinc Finger Protein Whose Expression Is Enhanced in a Neurodegenerative Disorder, Multiple System Atrophy (MSA). Genomics 54, 50–58 (1998).

32. Yabe, I. et al. Mutations in bassoon in individuals with familial and sporadic progressive supranuclear palsy-like syndrome. Scientific Reports 2018 8:1 8, 1–13 (2018).

33. De Lauzon-Guillain, B. et al. Mediation and modification of genetic susceptibility to obesity by eating behaviors. Am J Clin Nutr 106, 996–1004 (2017).

34. Backman, J. D. et al. Exome sequencing and analysis of 454,787 UK Biobank participants. Nature 2021 599:7886 599, 628–634 (2021).

35. Danecek, P. et al. Twelve years of SAMtools and BCFtools. Gigascience 10, 1–4 (2021).

36. McLaren, W. et al. The Ensembl Variant Effect Predictor. Genome Biol 17, 1–14 (2016).

37. Rentzsch, P., Witten, D., Cooper, G. M., Shendure, J. & Kircher, M. CADD: predicting the deleteriousness of variants throughout the human genome. Nucleic Acids Res 47, D886–D894 (2019).

38. Karczewski, K. J. et al. The mutational constraint spectrum quantified from variation in 141,456 humans. Nature 2020 581:7809 581, 434–443 (2020).

39. Morales, J. et al. A joint NCBI and EMBL-EBI transcript set for clinical genomics and research. Nature 2022 604:7905 604, 310–315 (2022).

40. Cingolani, P. et al. A program for annotating and predicting the effects of single nucleotide polymorphisms, SnpEff: SNPs in the genome of Drosophila melanogaster strain w1118; iso-2; iso-3. Fly (Austin) 6, 80–92 (2012).

41. Wang, Q. et al. Rare variant contribution to human disease in 281,104 UK Biobank exomes. Nature 2021 597:7877 597, 527–532 (2021).

42. Manichaikul, A. et al. Robust relationship inference in genome-wide association studies. Bioinformatics 26, 2867–2873 (2010).

43. Pedersen, B. S. & Quinlan, A. R. Who’s Who? Detecting and Resolving Sample Anomalies in Human DNA Sequencing Studies with Peddy. Am J Hum Genet 100, 406–413 (2017).

44. Auton, A. et al. A global reference for human genetic variation. Nature 2015 526:7571 526, 68–74 (2015).

45. Chen, S., et al. A genome-wide mutational constraint map quantified from variation in 76,156 human genomes. bioRxiv 2022.03.20.485034 (2022) doi:10.1101/2022.03.20.485034.

46. Viechtbauer, W. Conducting Meta-Analyses in R with the metafor Package. J Stat Softw 36, 1–48 (2010).

47. de Leeuw, C. A., Mooij, J. M., Heskes, T. & Posthuma, D. MAGMA: Generalized Gene-Set Analysis of GWAS Data. PLoS Comput Biol 11, e1004219 (2015).

48. Mak, T. S. H., Porsch, R. M., Choi, S. W., Zhou, X. & Sham, P. C. Polygenic scores via penalized regression on summary statistics. Genet Epidemiol 41, 469–480 (2017).

49. Pantazis, C. B. et al. A reference human induced pluripotent stem cell line for large-scale collaborative studies. Cell Stem Cell 29, 1685–1702.e22 (2022).

50. Santos, D. P., Kiskinis, E., Eggan, K. & Merkle, F. T. Comprehensive Protocols for CRISPR/Cas9-based Gene Editing in Human Pluripotent Stem Cells. Curr Protoc Stem Cell Biol 38, 5B.6.1–5B.6.60 (2016).

51. Skarnes, W. C., Pellegrino, E. & McDonough, J. A. Improving homology-directed repair efficiency in human stem cells. Methods 164–165, 18–28 (2019).

52. Xue, Y. et al. GenEditID: an open-access platform for the high-throughput identification of CRISPR edited cell clones. bioRxiv 657650 (2019) doi:10.1101/657650.

53. Merkle, F. T. et al. Generation of neuropeptidergic hypothalamic neurons from human pluripotent stem cells. Development 142, 633–643 (2015).

54. Kirwan, P., Jura, M. & Merkle, F. T. Generation and Characterization of Functional Human Hypothalamic Neurons. Curr Protoc Neurosci 81, 3.33.1–3.33.24 (2017).

55. Dowsett, G. K. C. et al. A survey of the mouse hindbrain in the fed and fasted states using single-nucleus RNA sequencing. Mol Metab 53, 101240 (2021).

56. Butler, A., Hoffman, P., Smibert, P., Papalexi, E. & Satija, R. Integrating single-cell transcriptomic data across different conditions, technologies, and species. Nature Biotechnology 2018 36:5 36, 411–420 (2018).

57. Hafemeister, C. & Satija, R. Normalization and variance stabilization of single-cell RNA-seq data using regularized negative binomial regression. Genome Biol 20, 1–15 (2019).

58. Love, M. I., Huber, W. & Anders, S. Moderated estimation of fold change and dispersion for RNA-seq data with DESeq2. Genome Biol 15, 1–21 (2014).

59. Zhou, Y. et al. Metascape provides a biologist-oriented resource for the analysis of systems-level datasets. Nature Communications 2019 10:110, 1–10 (2019).

