## Supplementary Informations for "Protein-truncating variants in *BSN* are associated with severe adult-onset obesity, type 2 diabetes and fatty liver disease"

### Supplementary Figures

**Figure S1 | Exome-wide rare (MAF <0.1%) variant associations with BMI.** (a) Combined Manhattan plot showing gene burden test results. Genes passing exome-wide significance (**P**$<1.33\times10^{-6}$) are highlighted. Points are annotated with variant predicted functional class (MISS REVEL; missense variants with REVEL scores (above 0.5 or 0.7), HC PTV; high confidence protein truncating variants). (b) Combined QQ plot for the gene burden tests.

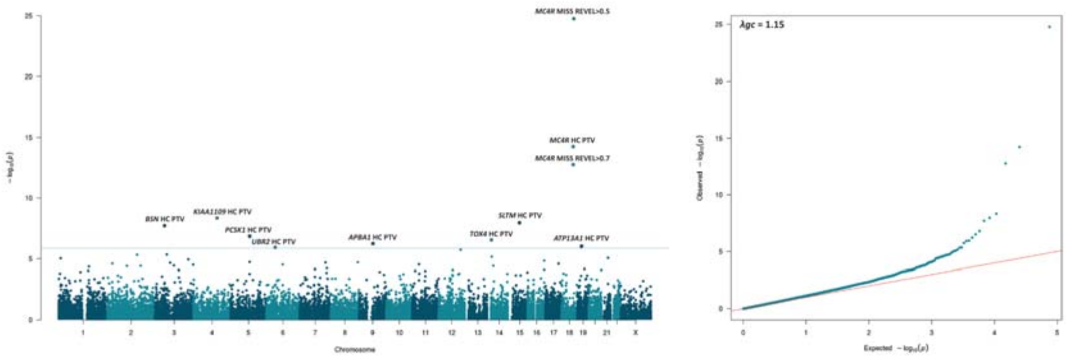

**Figure S2 | Synonymous variant associations with BMI.** Comparison of the QQ plots for gene burden associations for the three discovery variant mask and the synonymous variant mask.

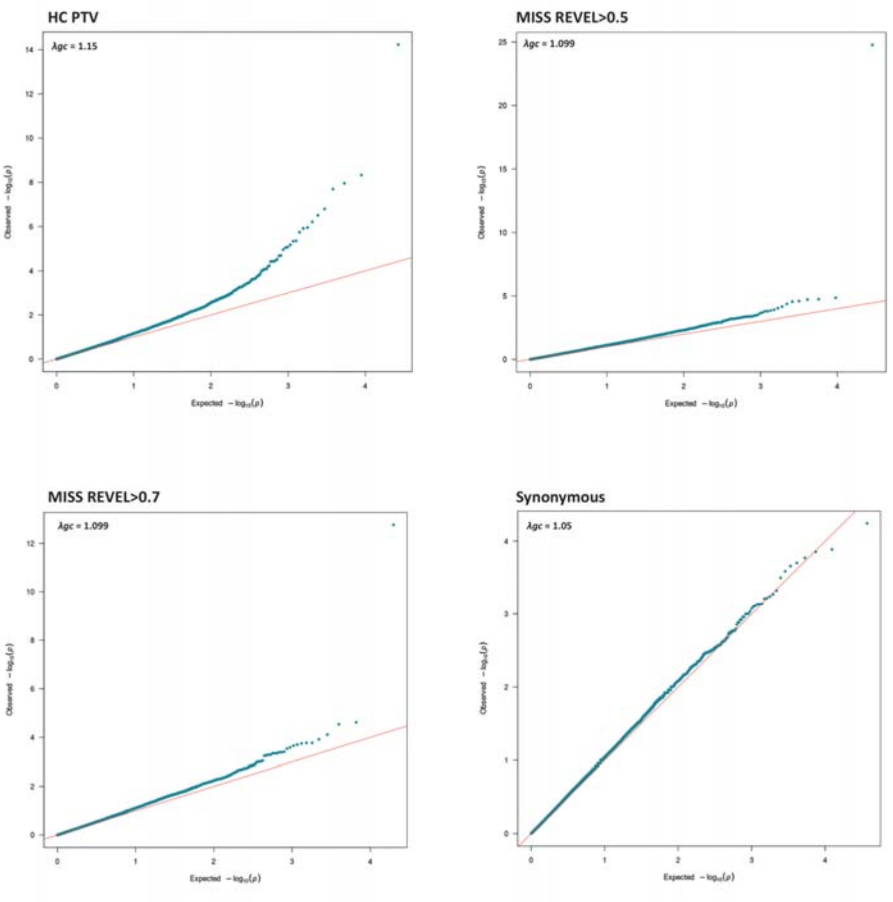

**Figure S3 | GWAS loci proximal to the exome-wide significant genes.** Associations in the UK Biobank BMI GWAS surrounding (±500kb) the 2 genes identified by exome-wide association with BMI, *BSN* (a) and *ATP13A1* (b). Relevant data are included in Table S2.

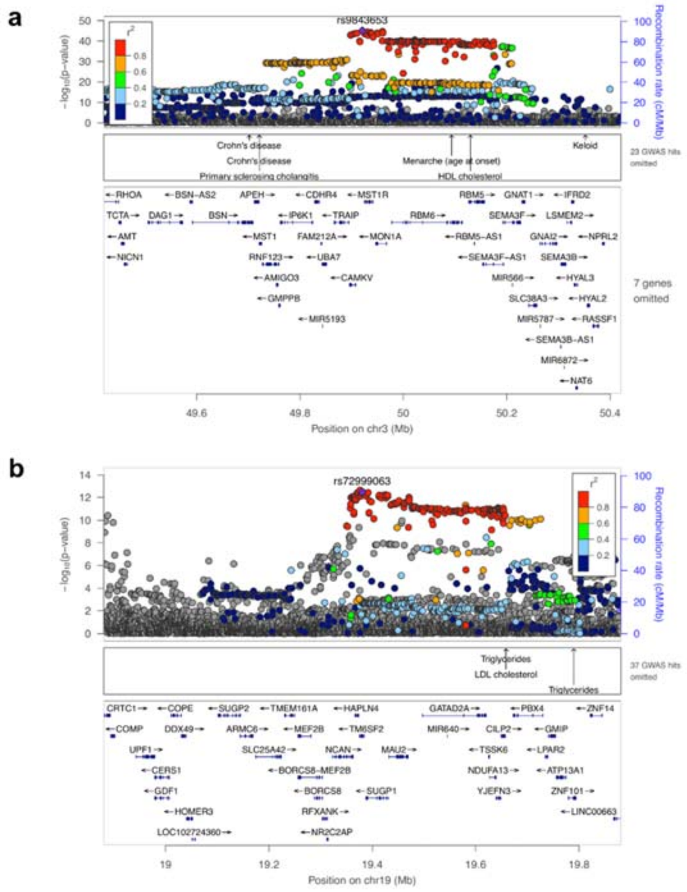

**Figure S4 | Replication of GWAS signals in Locke et al.**[1] Associations from the UK Biobank BMI GWAS at the *BSN* (a) and *ATP13A1* (b) loci, were further queried in an independent GWAS cohort. Relevant data are included in Table S2.

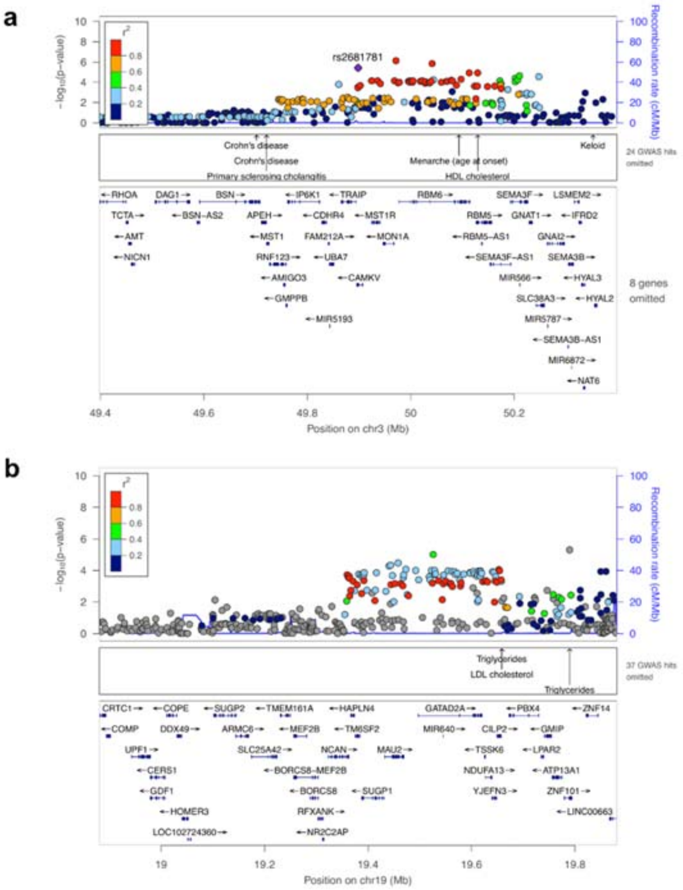

**Figure S5 | Geographical distribution of *APBA1* and *BSN* HC PTV carriers.**

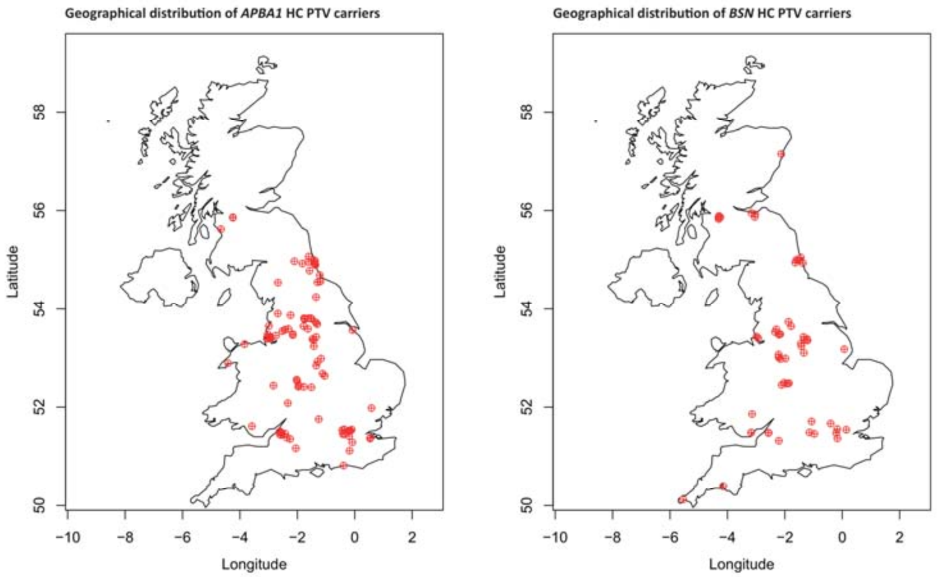

**Figure S6 | Interaction effect between PGS and *BSN* HC PTV carrier status.** Evidence of an interaction effect between PGS and carrier status of *BSN* HC PTV (beta=0.31, 95% CI: 0.12-0.60; p=0.01) such that the combined effect of one unit increase in PRS and being a carrier is greater than the sum of their marginal effect by 0.3 standard deviation in BMI.

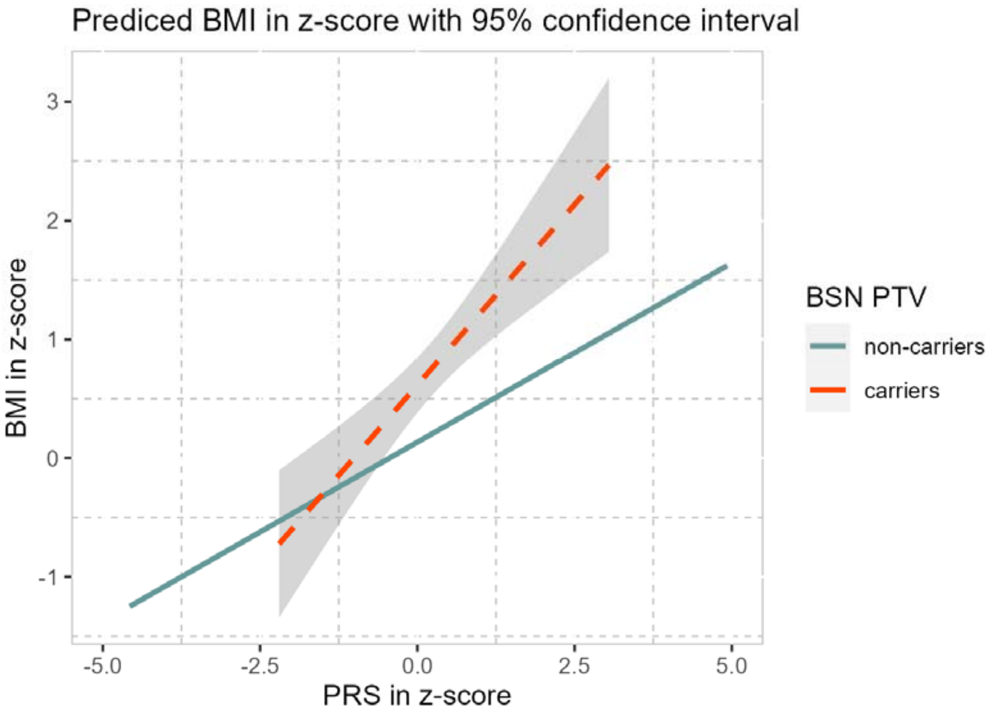

**Figure S7 | Kolf2 iPSCs differentiation into hypothalamic neurons.** Kolf2 iPSCs are shown here on day 36 of differentiation. The top panel shows the WT and bottom panel – *BSN* Het P399X.

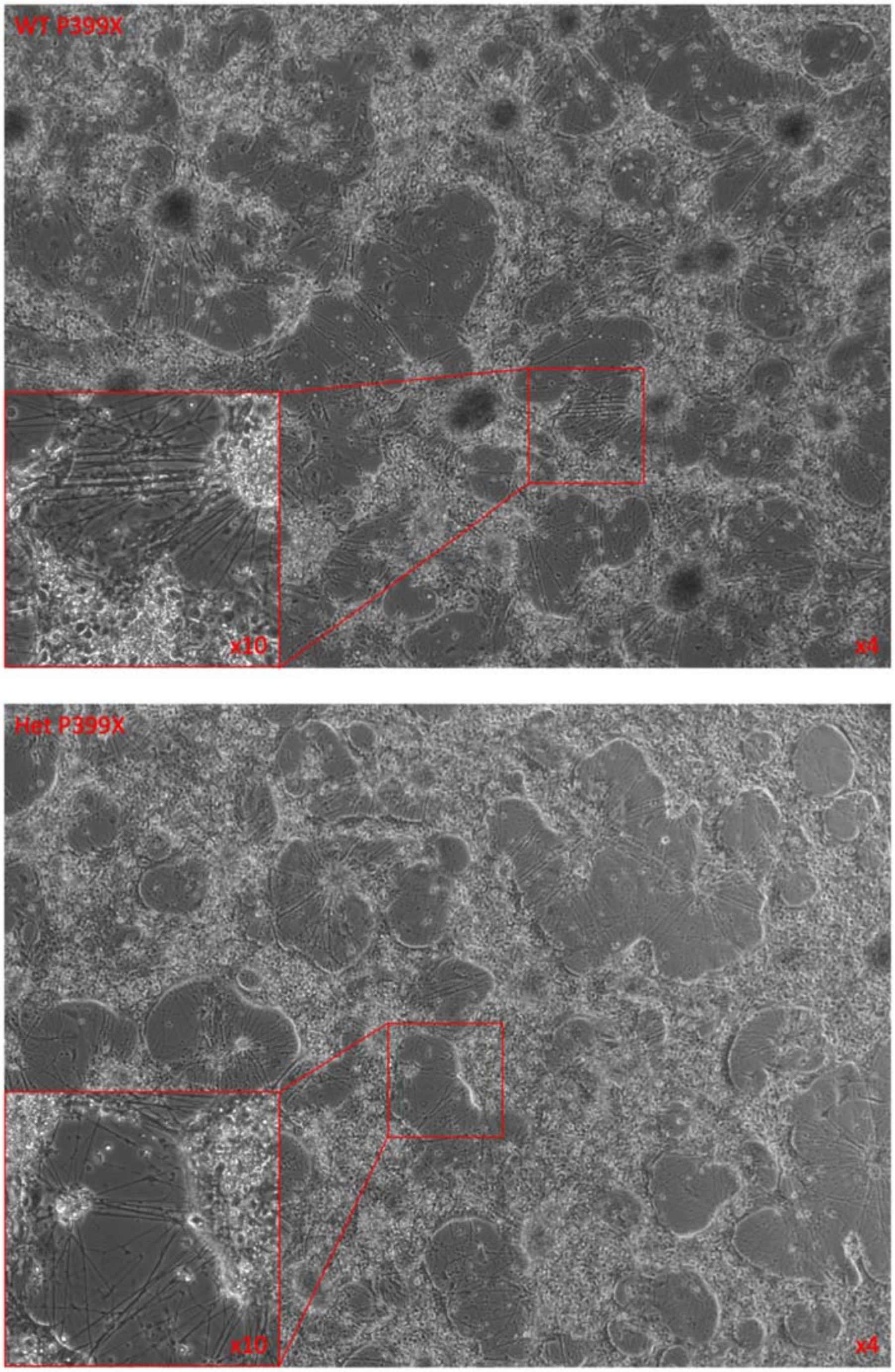

**Figure S8 | UMAP plots showing normalized expression of *RBFOX3*, *BSN* and *PCLO*.** Top row shows clustering of all the cells from single nucleus RNAseq. Bottom row shows re-clustering of clusters 1 and 5 from the top row.

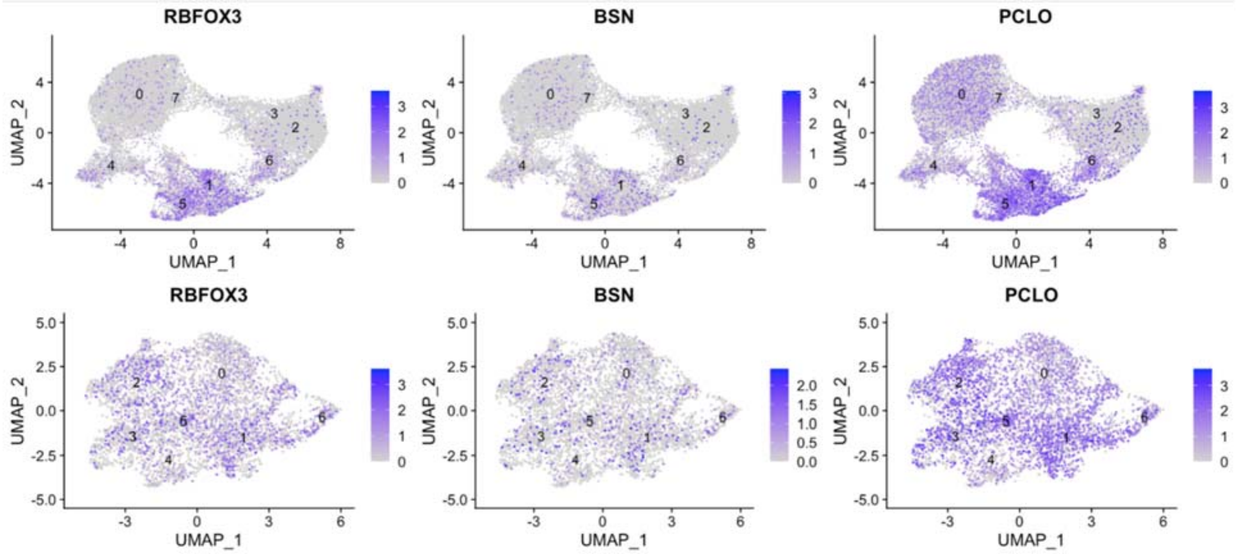

### References

1. Locke, A. E. *et al.* Genetic studies of body mass index yield new insights for obesity biology. *Nature 2015 518:7538* **518**, 197–206 (2015).
